# Detection of 2019 novel coronavirus in semen and testicular biopsy specimen of COVID-19 patients

**DOI:** 10.1101/2020.03.31.20042333

**Authors:** Ci Song, Yan Wang, Weiqin Li, Bicheng Hu, Guohua Chen, Ping Xia, Wei Wang, Chaojun Li, Zhibin Hu, Xiaoyu Yang, Bing Yao, Yun Liu

## Abstract

**Background:** As of March 11, 2020, the COVID-19 outbreak was declared as a pandemic. Expending our understanding of the transmission routes of the viral infection is crucial in controlling the outbreak. It is unclear whether the 2019 novel coronavirus (2019-nCoV) can directly infect the testes or male genital tract and be sexually transmitted from males.

**Methods:** From January 31 to March 14, 2020, 12 patients in recovery and one patient died of COVID-19 were included in this descriptive study. The clinical characteristics, laboratory findings, chest CT scans and outcome data were recorded. To examine whether there is sexual transmission from male, we employed realtime polymerase chain reaction testing (RT-PCR) to detect 2019-nCov in semen or testicular biopsy specimen.

**Findings:** The age range of the 12 patients in recovery was 22-38 years. None of the patients developed severe COVID-19 pneumonia. As of March 14, 2020, ten patients discharged from the hospital while the rest 2 had developed into recovery stage. All of the 12 patients in recovery tested negative for 2019-nCoV RNA in semen samples. Another patient aged 67 died in March 10, 2020, whose tissue sample via testicular biopsy was also tested negative for viral RNA.

**Conclusion:** No positive RT-PCR result was found in the semen or testicular biopsy specimen. The results from this study show no evidence of sexual transmission of 2019-nCov from males.

## Introduction

In the past two months, the total number of confirmed cases of COVID-19 in China has reached 80 000, and the global number has surpassed 300 000, more than 120 countries were affected [1]. Because of the speed and scale of transmission, the global COVID-19 outbreak was declared as a pandemic by the World Health Organization (WHO) on March 11, 2020 [2]. As reported [3], the 2019 novel coronavirus (2019-nCoV) can be efficiently transmitted between humans. Expending our understanding of the transmission routes of the viral infection is crucial in controlling the outbreak in the community.

Similar to severe acute respiratory syndrome (SARS) in 2003 and Middle East respiratory syndrome (MERS) in 2012, the COVID-19 infection causes a series of respiratory illness at visit, which indicates the virus most likely infects respiratory epithelial cells and spreads mainly via respiratory tract and contact transmission. In addition, the incidence of less common gastrointestinal manifestations like diarrhea, nausea and vomiting [4], along with the detection of virus in the feces specimens [5, 6], implies the viral gastrointestinal infection and fecal-oral transmission route. Recently, two studies have answered whether 2019-nCoV could spread sexually or vertically and pose risks to the fetus and neonate among female patients [7, 8]. No evidence was showed for sexual transmission or intrauterine infection caused by vertical transmission in women who develop COVID-19 pneumonia. However, according to the recent bioinformatic evidence [9, 10], the angiotensin-converting enzyme 2 (ACE2), a target for 2019-nCoV infection, is predominantly enriched in human testes, raising the urgent question of viral sexual transmission and paternal transmission route among males. Answer to this question is essential for formulating the principles of treatment and guidelines of sexual behavior when restored for men with 2019-nCoV infection.

Therefore, to explore the possibility of sexual transmission of this contagious virus from male, we examined the viral RNA in semen and testicular biopsy specimen respectively from 12 patients in recovery and one patients died in the acute phase of 2019-nCov infection.

## Methods

### Study design and patients

This study was approved by the Institutional Review Board (IRB) of Nanjing Medical University. In summary, we recruited 12 survived male COVID-19 patients from January 31 to March 14, 2020 in Wuhan. Patients with a severe COVID-19 pneumonia or severe underlying disease were not enrolled. Written informed consent was obtained from the 12 enrolled patient. In addition, one patient who had died of COVID-19 in March 10, 2020 in Wuhan was included, after we obtained written informed consent from his daughter. Totally, 13 patients were enrolled in this study. Diagnosis of COVID-19 pneumonia was based on the New Coronavirus Pneumonia Prevention and Control Program (7th edition) published by the National Health Commission of China [11]. COVID-19 pneumonia were diagnosed among suspected cases when nucleic acid tested positive by use of quantitative RT-PCR (qRT-PCR) on pharyngeal swab samples or the virus antibody (IgM or IgG) tested positive using colloidal gold test on serum.

### Data collection

We reviewed clinical characteristics, laboratory findings, chest CT scans and outcome data for all 13 men. All information was obtained and curated with a standardized data collection form. Two researchers also independently reviewed the data collection forms to double check the data.

For the 12 survived patients, semen samples were collected at recovery stage considering the patient’s physiological and psychological acceptance. Semen samples were obtained by masturbation according to the WHO guideline. To avoid virus contamination from non-semen sources, the process included passing urine, washing hands and penis with soap, drying hands and penis, and then ejaculating the semen into a sterile and wide-mouthed container [12]. Tissue sample was obtained from the dead patient with COVID-19 via directly testicular biopsy. Pharyngeal swab specimens were collected on admission day and every few days thereafter for the COVID-19 virus test (**Figure 1**). All samples were tested for 2019-nCoV by using qRT-PCR kits (Huirui biotechnology, Shanghai, China) recommended by the Chinese Center for Disease Control and Prevention (CDC) following WHO guidelines[11]. RT-PCR testing was used to detect 2019-nCov according to the recommended protocol. IgM and IgG antibodies in serum were measured by using a commercial colloidal gold test kit (Wondfo biotechnology, Guangzhou, China) recommended by the CDC. Sample collection, processing, and laboratory testing complied with WHO guidance. The recovery stage in this study was defined as period after the virus clearance (two continuous negatives of nucleic acid tests), or with the symptoms lessened and the chest CT showed a substantial resolution.

**Figure 1.**
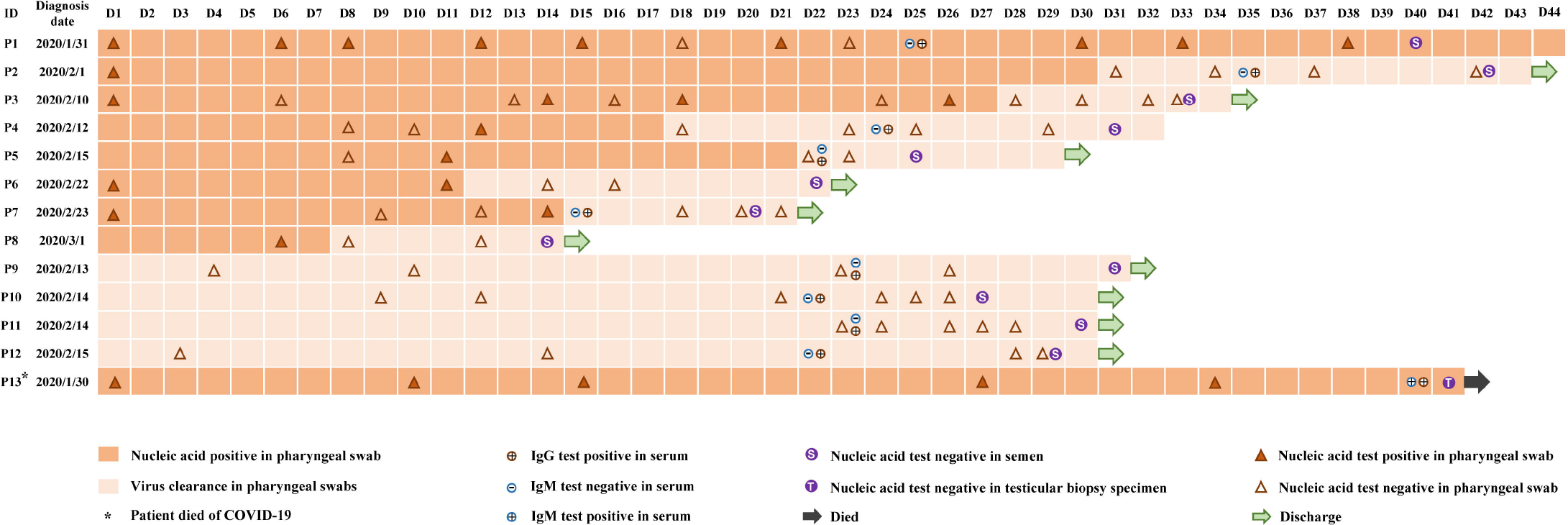
**Nucleic acid and antibody test among all specimens of the 13 patients during hospitalization. Date of diagnosis was defined as origin point (Day 1)**.

### Statistical analysis

Statistical analysis was done with STATA/SE 15.0. Continuous variables were expressed as a range. Categorical variables were expressed as number (%).

## Results

A total of 13 patients with COVID-19 were recruited in this study. The age range of the patients in recovery (Patient 1∼12) was 22-38 years; the dead patient (Patient 13) was 67 years old (**Table S1**). We collected the information of underlying diseases, including infection of Hepatitis B virus, Hepatitis C virus, tuberculosis and human immunodeficiency virus, diabetes, chronic hypertension, cardiovascular disease, cancer, COPD, sexually transmitted disease, prostatitis, epididymitis, and mumps orchitis. None of the patients had reported the above underlying diseases.

For the 12 patients in recovery (Patient 1∼12), none of them developed severe pneumonia. Most of them were categorized into mild/common subgroups (11 patients); one was categorized into asymptomatic subgroup. Seven patients presented with a fever during hospitalization, only one patient had a high fever with 39°C (Patient 11), other patients’ body temperatures fluctuated within a range of 36.5-38.5°C. Other symptoms of an upper respiratory tract infection were also observed: four patients had a cough, one patient had shortness of breath, and one reported a sore throat. Additionally, four patients had fatigue, one patient had a headache and one had a chest distress. Of the 12 patients, all of them underwent chest computed tomography (CT) on admission. The most common patterns on chest CT were patchy shadowing (5/12, 41.7%) and stripe shadows (3/12, 25.0%). Two (16.7%) patients showed ground-glass opacity in lungs. CT scans of the rest 2 (16.7%) patients showed normal images. All these patients were treated with antiviral therapy, 7 (58.3%) patients received antibiotic therapy. Three (25.0%) patients received intravenous corticosteroid therapy. Three (25%) patients received interferon therapy. One (8.3%) patient received immunoglobulin therapy. As of March 14, 2020, a total of 10 cases (83.3%) discharged from the hospital while the rest 2 had developed into recovery stage (Patient 4 has cleared the virus, while Patient 1 has the symptoms lessened and the recent chest CT showed a substantial resolution) and were kept in hospital for further observation. For the dead patient (Patient 13), he had a fever, cough, fatigue, shortness of breath, chest distress and dyspnea during hospitalization. The CT scans showed a patchy shadowing image. Except for the usual drugs, he was admitted to ICU in February 14, 2020 and received mechanical ventilation therapy. Unfortunately, the patient died at March 10, 2020.

Nucleic acid and antibody test among all specimens during hospitalization were recorded in **Figure 1**. Among the 12 patients in recovery, all of the 9 patients were tested for IgM negative and IgG positive. Eight of them have cleared the virus when the IgG turned positive, while only one patient (Patient 1) kept nucleic acid positive on pharyngeal swab specimens. Four patients (Patient 9, 10, 11 and 12) kept nucleic acid negative but with IgG positive during hospitalization. The dead patient (Patient 13) was tested for IgM positive and IgG positive, while positive for 2019-nCoV RNA in pharyngeal swab, indicating an acute phase of infection.

Patient 2-11 tested negative for 2019-nCoV RNA in semen samples when the virus has been cleared in pharyngeal swabs. Patient 1 has also developed into recovery stage and tested negative for 2019-nCoV RNA in semen sample, but still kept viral RNA positive in pharyngeal swab. Patient 13, still in the acute phase, and was tested negative for 2019-nCoV RNA in the tissue sample via testicular biopsy. In summary, all of the patients were tested negative for 2019-nCoV RNA in semen samples.

## Discussion

The route of transmission of 2019-nCov is worth exploring to control the virus spread. Respiratory droplets and intimate contact are the two main transmission routes of 2019-nCov [13, 14]; evidence for fecal-oral transmission route was provided [15, 16]; vertical and sexual transmissions among female patients have not been recognized [7, 8]. Here, we collected semen samples from 12 patients in recovery, and the semen samples were tested negative for 2019-nCov RNA through PCR assay. Absence of viral RNA in testicular biopsy tissue further indicated that the virus would not directly infect the testes or male genital tract even in the acute phase. No evidence showed that 2019-nCov could be sexually transmitted from males.

Globally, there are more than 20 recognized sexually transmitted infections (STIs) [17, 18], some are viruses including HIV [19], HSV [20], Zika virus [21], Ebola virus [22] and so on. Most of them are capable of causing viremia and found in human semen [18]. Our results showed that the 2019-nCov could not be found in semen. Although positive PCR test of 2019-nCov in blood samples was reported, the percentage was limited (1%) [6], which suggested limited possibilities for viremia onset.

Numerous studies have revealed the role of cell surface ACE2 as the cellular receptor for SARS-Cov spike protein [23, 24]. ACE2 has also been proven to be a major receptor of the novel 2019-nCoV due to it shares high sequence (share ∼76% amino acid identity) similarity with SARS-Cov [25]. Also, recent studies have found that ACE2 is highly expressed in spermatogonia, Leydig and Sertoli cells in the human testis by single-cell transcriptome and bioinformatics analysis, and speculated that human testes are a potential target of 2019-nCov infection [9, 10]. However, consistent with our study, the previous systematical study reported that the cells in testis cannot be infected by SARS-CoV despite the expression of high levels of ACE2 [26]. On the other hand, although few studies previously reported orchitis as a complication of SARS [27], which mostly due to the continuous high fever and virus-induced autoimmune response among severe patients. Hence, the presence of orchitis among SARS-infected patients could not be concluded as the virus directly targeted the testes. Further mechanism investigation of the potential virus attacks on the testes is warranted.

Our study has several limitations. First, the sample size was relatively small in which only 13 patients with 2019-nCov positive was included; more patients are needed to confirm our findings in the future. Second, multiple nucleic acid tests on semen should be taken during the course of disease, especially in the incubation period. However, multiple collection of semen sample was difficult. Nevertheless, this is the first study that directly evidenced the absence of 2019-nCov in semen and testicular biopsy specimen of male COVID-19 patients, which indicated that 2019-nCov can not straightway infect the testes and male genital tract.

## Data Availability

We obtained the data from Wuhan No.1 Hospital and Jinling Hospital. The ownership of the data belongs to the two hospitals. Researchers who meet the criteria for access to confidential data can contact the corresponding authors to request the data.

## Acknowledgement

This study was funded in part by the National Key Research and Development Program of China (2016YFC1000200, 2018YFC1004200, 2018YFC1004700); the State Key Program of National Natural Science of China (31530047); the National Natural Science Foundation of China (81903382, 81971374), the Natural Science Foundation of Jiangsu Province (BK20190652), China Postdoctoral Science Foundation (General Progaram, 2019M651900). Written informed consent was obtained from each enrolled patient.

## Author Approval

All authors have seen and approved the manuscript.

## Data sharing

Researchers who meet the criteria for access to confidential data can contact the corresponding authors to request the data.

## Ethical approval

This study was reviewed and approved by the Institutional Review Board (IRB) of Nanjing Medical University (approval number 2020-008).

## Author’s contribution

Zhibin Hu, Xiaoyu Yang, Bing Yao and Yun Liu made substantial contributions to the study concept and design. Yan Wang, Weiqin Li, Bing Yao, and Yun Liu took responsibility for the acquisition and check of data. Zhibin Hu, Xiaoyu Yang, Ci Song and Xiaoyu Yang took responsibility for analysis and interpretation of the data; Bicheng Hu and Bing Yao were responsible for laboratory detection; Ci Song and Xiaoyu Yang were in charge of the manuscript draft. Ci Song took responsibility for the statistical analysis; Guohua Chen, Ping Xia, Wei Wang provided administrative, technical, and material support; Zhibin Hu, Xiaoyu Yang, Bing Yao, and Yun Liu made substantial revisions to the manuscript.

## Reference

1. World Health Organization. Coronavirus disease 2019 (COVID-19) Situation Report – 63. Available at: https://www.who.int/docs/default-source/coronaviruse/situation-reports/20200323-sitrep-63-covid-19.pdf?sfvrsn=d97cb6dd_2. Accessed 23 March 2020.

2. Ghebreyesus TA. WHO Director-General’s opening remarks at the media briefing on COVID-19 - 23 March 2020. Available at: https://www.who.int/dg/speeches/detail/who-director-general-s-opening-remarks-at-the-media-briefing-on-covid-19---23-march-2020. Accessed 23 March 2020.

3. Chan JF, Yuan S, Kok KH, et al. A familial cluster of pneumonia associated with the 2019 novel coronavirus indicating person-to-person transmission: a study of a family cluster. Lancet (London, England) 2020; 395(10223): 514–23.

4. Guan WJ, Ni ZY, Hu Y, et al. Clinical Characteristics of Coronavirus Disease 2019 in China. The New England journal of medicine 2020. Available at: https://www.nejm.org/doi/pdf/10.1056/NEJMoa2002032?articleTools=true. Accessed 28 February, 2020.

5. Peng L, Liu J, Xu W, et al. 2019 Novel Coronavirus can be detected in urine, blood, anal swabs and oropharyngeal swabs samples. Available at: https://www.medrxiv.org/content/10.1101/2020.02.21.20026179v1. Accessed 25 February, 2020.

6. Wang W, Xu Y, Gao R, et al. Detection of SARS-CoV-2 in Different Types of Clinical Specimens. Jama 2020. Available at: https://jamanetwork.com/journals/jama/fullarticle/2762997. Accessed 11 March, 2020.

7. Cui P, Chen Z, Wang T, et al. Clinical features and sexual transmission potential of SARS-CoV-2 infected female patients: a descriptive study in Wuhan, China. Available at: https://www.medrxiv.org/content/10.1101/2020.02.26.20028225v2. Accessed 3 March, 2020.

8. Chen H, Guo J, Wang C, et al. Clinical characteristics and intrauterine vertical transmission potential of COVID-19 infection in nine pregnant women: a retrospective review of medical records. Lancet (London, England) 2020; 395(10226): 809–15.

9. Wang Z, Xu X. scRNA-seq Profiling of Human Testes Reveals the Presence of ACE2 Receptor, a Target for SARS-CoV-2 Infection, in Spermatogonia, Leydig and Sertoli Cells. Available at: https://www.preprints.org/manuscript/202002.0299/v1. Accessed 21 February 2020.

10. Fan C, Li K, Ding YH, et al. ACE2 Expression in Kidney and Testis May Cause Kidney and Testis Damage After 2019-nCoV Infection. Available at: https://www.medrxiv.org/content/10.1101/2020.02.12.20022418v1. Accessed 13 February, 2020.

11. National Health Commission of the People’s Republic of China. New coronavirus pneumonia prevention and control program (7th edn). Available at: http://www.nhc.gov.cn/yzygj/s7653p/202003/46c9294a7dfe4cef80dc7f5912eb1989.shtml. Accessed 4 March, 2020.

12. World Health Organization, Department of Reproductive Health and Research. WHO laboratory manual for the examination and processing of human semen Fifth edition. Available at: https://www.who.int/reproductivehealth/publications/infertility/9789241547789/en/. Accessed 2010.

13. Huang C, Wang Y, Li X, et al. Clinical features of patients infected with 2019 novel coronavirus in Wuhan, China. Lancet (London, England) 2020; 395(10223): 497–506.

14. Chen N, Zhou M, Dong X, et al. Epidemiological and clinical characteristics of 99 cases of 2019 novel coronavirus pneumonia in Wuhan, China: a descriptive study. Lancet (London, England) 2020; 395(10223): 507–13.

15. Xiao F, Tang M, Zheng X, et al. Evidence for gastrointestinal infection of SARS-CoV-2. Gastroenterology 2020. Available at: https://www.gastrojournal.org/article/S0016-5085(20)30282-1/fulltext. Accessed 3 March, 2020.

16. Kanne JP. Chest CT Findings in 2019 Novel Coronavirus (2019-nCoV) Infections from Wuhan, China: Key Points for the Radiologist. Radiology 2020; 295(1): 16–7.

17. Bernstein K, Bowen VB, Kim CR, et al. Re-emerging and newly recognized sexually transmitted infections: Can prior experiences shed light on future identification and control? PLoS medicine 2017; 14(12): e1002474.

18. Salam AP, Horby PW. The Breadth of Viruses in Human Semen. Emerging infectious diseases 2017; 23(11): 1922–4.

19. Rambaut A, Posada D, Crandall KA, Holmes EC. The causes and consequences of HIV evolution. Nature reviews Genetics 2004; 5(1): 52–61.

20. Patton ME, Bernstein K, Liu G, Zaidi A, Markowitz LE. Seroprevalence of Herpes Simplex Virus Types 1 and 2 Among Pregnant Women and Sexually Active, Nonpregnant Women in the United States. Clinical infectious diseases : an official publication of the Infectious Diseases Society of America 2018; 67(10): 1535–42.

21. Foy BD, Kobylinski KC, Chilson Foy JL, et al. Probable non-vector-borne transmission of Zika virus, Colorado, USA. Emerging infectious diseases 2011; 17(5): 880–2.

22. Barre-Sinoussi F, Chermann JC, Rey F, et al. Isolation of a T-lymphotropic retrovirus from a patient at risk for acquired immune deficiency syndrome (AIDS). 1983. Revista de investigacion clinica; organo del Hospital de Enfermedades de la Nutricion 2004; 56(2): 126–9.

23. Li W, Sui J, Huang IC, et al. The S proteins of human coronavirus NL63 and severe acute respiratory syndrome coronavirus bind overlapping regions of ACE2. Virology 2007; 367(2): 367–74.

24. Wu K, Li W, Peng G, Li F. Crystal structure of NL63 respiratory coronavirus receptor-binding domain complexed with its human receptor. Proceedings of the National Academy of Sciences of the United States of America 2009; 106(47): 19970–4.

25. Hoffmann M, Kleine-Weber H, Krüger N, et al. The novel coronavirus 2019 (2019-nCoV) uses the SARS-coronavirus receptor ACE2 and the cellular protease TMPRSS2 for entry into target cells. Available at: https://www.biorxiv.org/content/10.1101/2020.01.31.929042v1. Accessed 31 January, 2020.

26. Ding Y, He L, Zhang Q, et al. Organ distribution of severe acute respiratory syndrome (SARS) associated coronavirus (SARS-CoV) in SARS patients: implications for pathogenesis and virus transmission pathways. The Journal of pathology 2004; 203(2): 622–30.

27. Lardhi AA. Henoch-Schonlein purpura in children from the eastern province of Saudi Arabia. Saudi medical journal 2012; 33(9): 973–8.

